# “That’s part of the culture:” A peer-to-peer study of reporting student mistreatment

**DOI:** 10.1101/2024.01.02.23300029

**Authors:** Alissa S. Chen, Bernice Yau, Kelsey B. Montgomery, Nicole Dubuque, Dana McDowelle, David Berg, Stephen R. Holt

**Affiliations:** Department of Internal Medicine, Yale School of Medicine, New Haven, Connecticut, USA; Department of Psychiatry, Columbia University Medical Center, New York, New York, USA; Department of Surgery, University of Alabama at Birmingham, Birmingham, Alabama, USA; McGovern Medical School, University of Texas Health Science Center, Houston, Texas, USA; Department of Psychiatry, Yale School of Medicine, New Haven, Connecticut, USA

**Keywords:** learning environment, undergraduate medical education, student mistreatment, professionalism

## Abstract

**Introduction:** Forty percent of graduating medical students report experiencing student mistreatment; however, most cases go unreported.

**Methods:** Peer-to-peer, semi-structured interviews with current medical students were conducted to understand medical students’ experiences with mistreatment and reporting. These interviews were inductively coded, and themes were identified to elucidate reasons students do and do not report mistreatment.

**Results:** Twenty-one students were interviewed who described a total of 34 mistreatment incidents. Four main groups of factors that students consider when deciding to report mistreatment were identified: personal, situational, structural, and climate. Personal factors were intrinsically tied to the participant, including their feelings or concerns about mistreatment. Situational factors related to the act of mistreatment, such as who the perpetrator was or the actions of bystanders. Structural factors included elements of the reporting system. Climate factors were concerns related to how the institution viewed mistreatment and the student’s place in medical education.

**Discussion:** This peer-to-peer study revealed four main groups of factors, all of which are influenced by the culture of the institution. Participants were impacted by the inactions of witnesses and their personal sense of justice. An approach to facilitating reporting of student mistreatment must be grounded in improving the culture of medical education.

## Introduction

Despite being described as early as the 1980’s, student mistreatment continues to be a challenge in medical education (Mavis et al. 2014; Silver and Glicken 1990). According to the most recent American Academy of Medical Colleges (AAMC) Graduation Questionnaire (GQ), the prevalence of mistreatment in graduating medical students is 38.12% (AAMC 2023). Surveys in Asia, Europe, and Canada have found similar results (Ahmer et al. 2008; Alzahrani 2012; Gágyor 2012; Moscarello et al. 1994).

Studies have shown that mistreatment has deleterious psychological effects on students. Students who experience mistreatment are more likely to show symptoms of anxiety, depression, post-traumatic stress, and hostility (Haglund et al. 2009; Heru et al. 2009; Richman et al. 1992). Mistreated students are also likely to experience burnout, which has been strongly linked with a decrease in empathy (Cook et al. 2014; Hariharan and Griffin 2019; Neumann et al. 2011).

Moreover, student mistreatment has led students to question their career choices. In a survey of 2,316 students at 16 different medical schools (Frank et al. 2006), researchers found that students who experienced mistreatment were more likely to regret becoming a doctor. In particular, students who previously planned on pursuing a career in academic medicine were less likely to plan on doing so after being mistreated (Haviland et al. 2011).

Regrettably, a roadblock to preventing student mistreatment is a lack of reporting. In the most recent GQ, while nearly 40% of respondents noted they were mistreated, only 26.9% of those respondents stated they had reported mistreatment (AAMC 2023). The GQ lists several factors for respondents to elucidate why they chose not to report. The most common factors were “the incident did not seem important enough to report” (51.0%), followed by “I did not think anything would be done about it” (41.7%), and “fear of reprisal” (32.1%) (AAMC 2023). However, this information merely scratches the surface of why students do not report mistreatment.

A few qualitative studies have begun to further elucidate the barriers to reporting (Bell et al. 2021; Chung et al. 2018; Colenbrander et al. 2020). In an interview study by Bell et al. (2021), researchers described the process a medical student takes while deciding to report mistreatment. Throughout the journey the medical student takes, their decision is mainly influenced by their experience with their institution. The researchers describe one of the steps as “situating,” where medical students understand their place in the medical hierarchy to understand why they were mistreated and what they should do about it. Two additional qualitative studies found that medical students perceived the culture of an institution as a barrier to reporting (Chung et al. 2018; Colenbrander et al. 2020). Students mentioned that the hierarchy of medicine prevented them from reporting, but not reporting also prevented culture change (Colenbrander et al. 2020). Bell et al. (2021) found in particular that medical students saw the inaction of bystanders impactful. Passive bystanders seemed to normalize the mistreatment they experience, making the situation more distressing. Given the power of the hierarchy in medicine to prevent reporting, bystanders who are superior to either the medical student or the perpetrator may play a key role in promoting reporting of mistreatment.

While the qualitative studies available in the literature have expanded upon perceived barriers to reporting mistreatment, it is not clear what factors motivate medical students to report mistreatment. By understanding what motivates students to report, we can more effectively improve the process for reporting. In this study, we conducted peer-to-peer, semi-structured interviews with current medical students who had experienced mistreatment to understand what factors they considered when deciding to report. Studies have shown that peer-to-peer, or insider, interviews can allow interviewees to divulge more information than they would to an outsider because of increased confidence in the interviewer (Byrne et al. 2015; Hockey 1993). One-on-one interviews with peers, rather than non-peer led interviews or focus groups, may provide even more information, in light of the sensitive nature of the topic.

## Methods

The University of Texas Health Science Center Institutional Review Board approved the study protocol. We conducted a single-site, qualitative study using in-depth key informant interviews of current medical students.

### Context

In the United States, undergraduate medical education lasts four years. The first two years are typically classroom based, and the second two years are clinically based. When doing clinical duties, students may work in the inpatient wards, outpatient clinic, and surgical areas. They may be supervised by residents or attending physicians.

### Recruitment

The study institution was a large, urban academic institution in the southern United States. Students were recruited via emails sent in April 2019 by student class presidents to the first through fourth year classes’ listservs, which reached approximately 1,000 students. Students were eligible to participate if they were current MD candidates and had experienced mistreatment during medical school, as defined by them. The recruitment email contained the study description, the time required, and the risks and benefits. It also explained that the interviews would be conducted by student researchers and transcribed by students. Students were informed that participation did not equate to reporting their mistreatment incident. Interested students emailed the student researchers, who then provided information for scheduling their interviews. All interested students were included in the study.

### Interviews

Interviews were conducted with one participant and two peer researchers: one third year medical student (KM) and one fourth year medical student (AC). A total of 21 interviews were conducted between April-May 2019. Interviews were 15 to 45 minutes in length. Prior to the initiation of the interview, participants were asked to review the letter of information before providing verbal consent. The interviewers utilized a list of open-ended questions developed by the study authors that fit the research aims. The categories of questions asked included the student’s personal experience with mistreatment, their experience with reporting, their reasons for choosing whether or not to report, their general thoughts about the reporting process, and any suggestions for improvement of reporting. If the participant experienced more than one episode of mistreatment, they were asked to describe each instance with the same set of questions. If further clarification was necessary to fully understand a participant’s answer, the interviewers would ask follow-up questions.

Interviews were recorded with a digital audio recorder. Audio files were stored on a password protected database only accessible to student researchers. After completion of the interviews, audio files were transcribed by student researchers and student study authors. Any identifying information (i.e. student, faculty, staff, resident, or fellow name, locations, specialties) was redacted during transcription and prior to data analysis.

### Data Analysis

Coding of the transcripts was performed based on grounded theory and inductive analysis described by Patton (2002). Two researchers (AC and BY) reviewed all transcripts prior to code creation. They then used inductive analysis to code three transcripts exhaustively for mistreatment characteristics (e.g., persons involved, type of mistreatment, setting), students’ responses to mistreatment, and reasons for reporting or not reporting until no new codes emerged. These codes were compared and rectified via consensus in order to create an initial code book. After developing a code book, the two researchers applied the codes to the rest of the transcripts independently. Codes of the remaining transcripts were then compared and rectified. After open coding was completed, axial coding of the code book was completed to categorize individual codes into groups. Code groups were then analyzed to identify overarching themes based on similarities in concepts described until no new themes emerged (i.e., until theoretical saturation was reached). These themes are presented in the results section.

## Results

### Descriptive data

Interviews were conducted with 21 participants, of whom 13 identified as women and 8 identified as men (**Table 1**). The median age of participants was 25 years (mean = 26.43, SD = 3.32).

**Table 1.** Descriptive Statistics.

| Characteristic | N |
| --- | --- |
| Gender |  |
| Man | 8 |
| Woman | 13 |
| Race |  |
| White | 10 |
| Asian | 6 |
| Hispanic, Latino, or Spanish | 3 |
| Black or African American | 1 |
| American Indian or Alaska Native | 1 |
| Medical School Year |  |
| Second | 1 |
| Third | 9 |
| Fourth | 11 |

Thirty-four incidents of mistreatment were described by the 21 participants. Of these incidents, 12 were reported (using online reporting software, discussing with student affairs, or discussing with the course director) and 22 were not reported. Participants described an average of 1.6 mistreatment incidents each (minimum = 1, maximum = 3).

Through inductive analysis of the responses students had to mistreatment and their reasons for reporting and not reporting, four overarching groups of factors that students consider when deciding whether or not to report mistreatment emerged: personal, situational, structural, and climate. **Figure 1** demonstrates each reason for and against reporting within each factor.

**Figure 1:**
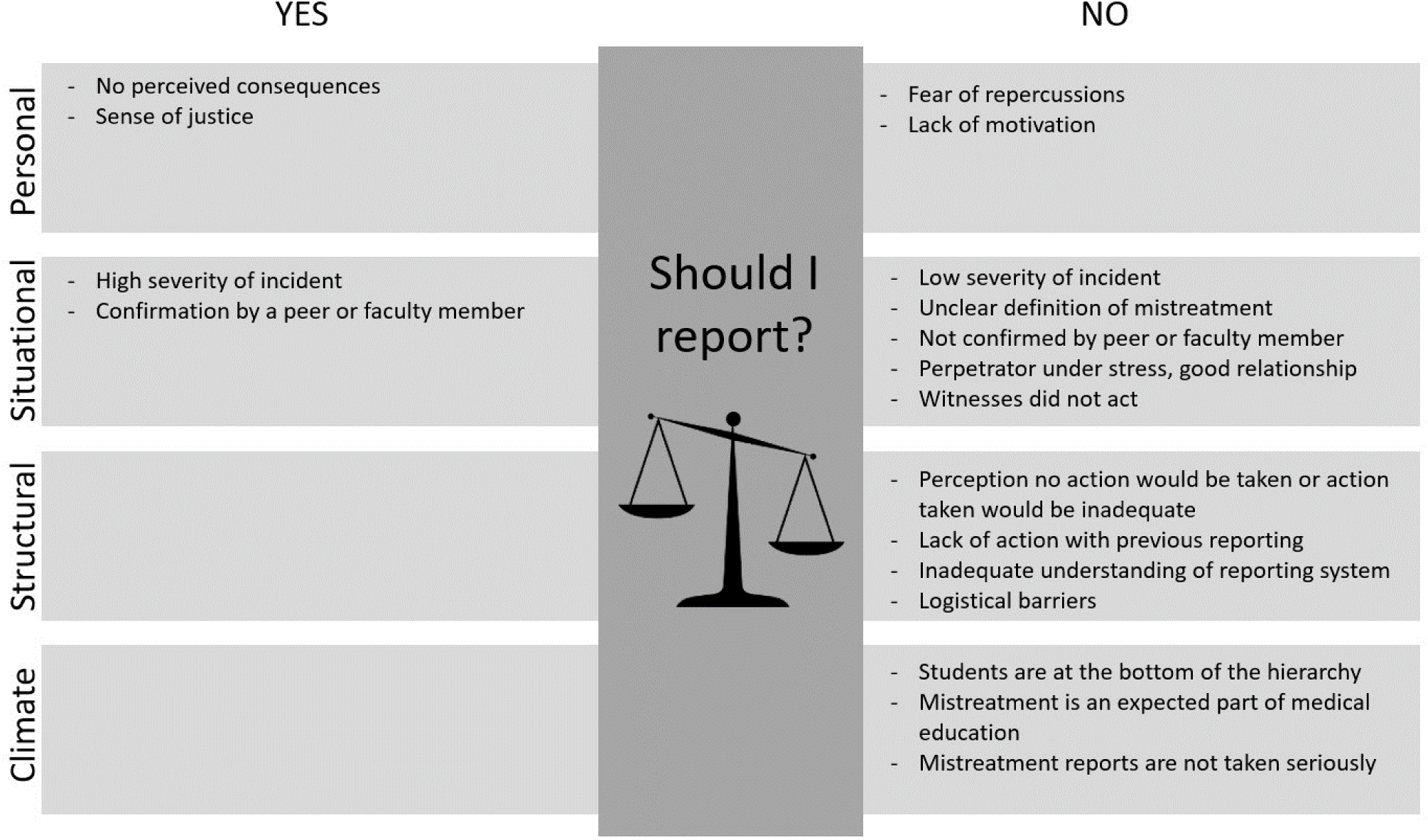
Factors that impact reporting of student mistreatment.

### Personal factors: “What are going to be the consequences to me?”

We defined personal factors as factors which were intrinsically related to the participant, such as feelings or concerns about the mistreatment or their chosen career path. Multiple personal factors contributed to a student’s willingness to report, including concern for repercussions, sense of justice, and lack of personal motivation. The main deterrent to reporting was concern for repercussions. This includes fear of retaliation, which could be in the form of impairing grades or advancement. Students noted that their grades or their careers were more important to them than ensuring action was taken on their incident.

> *I know that they take, like, these kinds of reports seriously no matter what it is, but at the same time it’s like, what are going to be the consequences to me?* -Participant R

However, this was not always a deterrent. Some students stated that they were not going into the same specialty as the perpetrator, and they felt safe reporting as they did not fear repercussions on their career. Others already had their grade finalized and thus felt confident in reporting.

> *I think it was a little bit easier for me personally to report it because I knew I wasn’t going into [that specialty]. And if I had to do further rotations in this same environment, I don’t know how comfortable I would have felt going through that same process.* -Participant A

Another personal factor was a sense of justice. Students felt that they needed to protect other students or that they felt a duty to report. For some, this duty originated from a need they felt for corrective action. This sense of justice could also be a deterrent to reporting as some students perceived that action had already been taken, and it was not necessary for them to report their incident.

> *I knew that if I knew a female or minority student that took the rotation behind me and I hadn’t said anything, I would feel responsible for that student’s potentially poor experience. So at that point, it solidified that I needed to bring it to someone’s attention.* -Participant J

Finally, some students lacked personal motivation to report, which had multiple etiologies. For some, this stemmed from their assessment of nothing to gain from reporting.

> *I kind of looked at that as I’m not going to gain anything out of reporting this… close the chapter and put it behind me rather than pursue a further headache.* – Participant J

### Situational Factors: “I felt like that was something that definitely crossed the line”

Situational factors were defined as attributes of the act of mistreatment such as who the perpetrator was, where the mistreatment took place, and what type of mistreatment it was. The type and severity of the mistreatment was weighed by many students in this study. Students struggled with the ambiguous definition of mistreatment. If they were uncertain if they experienced mistreatment, they were less likely to report. Many noted either that the incident was either not severe enough to report, or the incident was so severe it required reporting. More specifically, mistreatment based on race was much more likely to be viewed severe enough to compel students to report.

> *When the wording came into race, I felt like that was something that definitely crossed the line and that was unacceptable, no matter what work environment it was.* -Participant A

Participants often relied on the opinions of confidants to determine if they should report an incident. Students also felt more comfortable reporting if their incident confirmed by confidants, namely friends or family.

> *It didn’t even occur to me that it might be some form of mistreatment like until months later when I was just telling somebody else about it…They were like “oh yeah, that’s horrible. You should have told someone about that.”* -Participant B

Additionally, participants described the actions of bystanders in their assessment. Parties varied from a single bystander to large groups of bystanders present. Some were peers of the student or the perpetrator, and others were the superiors of the perpetrator or student. Bystanders described by the participants did not intervene. Some of them gave non-verbal cues, which participants interpreted as an acknowledgement of the event.

> *The attending turned towards me and he was like “what the f*** are you laughing at medical student?” … I stopped laughing immediately. The resident kind of gave me an “I’m sorry” glance.* -Participant D

Largely, however, most bystanders did not take notice of the event and might have said they “didn’t hear” (Participant T) what happened if confronted by the participant. Participants who noted the inaction of bystanders were less likely to report mistreatment.

> *“There are three other residents in the room with her, one of them was actually above her… the chief was in there also, and no one said anything and I’m like, this is ridiculous”* – Participant S

Finally, the victim’s assessment of the perpetrator had an impact on their willingness to report. For instance, if the participant thought the perpetrator was under stress or did not intend to mistreat them, they felt less inclined to report. Others felt they had a good relationship with the perpetrator or the perpetrator h ad other redeeming qualities, and they did not want them to be punished.

> *I thought he was a nice guy. I think he maybe really had poor taste in that moment, and so I didn’t want to get him in trouble.* -Participant N

### Structural factors: “I didn’t think that really anything would come of it.”

We defined structural factors as factors related to the reporting apparatus available to students. This included the reporting software, designated reporting faculty, action taken as a result of the report, and communication with the student on the outcome of the report. One of the common themes was that students felt that if they did report, no action would be taken. They felt that they would not be taken seriously and that the administration may not agree this was mistreatment. For some, this was based on prior poor experience with reporting. For others, they felt that there were no long-term consequences that the administration could take that would equal the severity of the event. One student who was mistreated twice, stated:

> *So the primary reason is that I didn’t think that really anything would come of it because I felt like the first [mistreatment incident] was so much more serious, and nothing came of that reporting.* -Participant F

Additionally, multiple logistical barriers were noted by students. Some were uncertain of where they should report. Others did not understand the process and outcomes of reporting (i.e. where the data goes, what happens with the data, when action is taken). Many participants noted they never received information on what happens with reports and thus did not feel any action would be taken. As a result, they felt reporting was futile.

> *I think the biggest thing really is the fact that we don’t know what happens with reporting. Like we trust the system works but we don’t have a way of knowing it works essentially, and I think that leads us to not reporting as well.* -Participant O

### Climate factors: “I just expected some degree of abuse”

Climate factors were participants’ perceptions of the culture of their medical institution and medical education. The word “climate” was used to describe this factor as it describes students’ perception of the culture. We will use the word “culture” to describe the beliefs and values within an institution; these terms have also been used in safety literature (Mearns and Flin 1999). Many students expressed that the culture of medicine is hierarchical. This lends itself not only to a high incidence of mistreatment but also to the perception by faculty and administrators that mitigating student mistreatment is not a priority. Students expressed that they felt that as the person at the bottom of the hierarchy, it was expected that they would be mistreated.

> *Honestly, on some level, I just expected some degree of abuse to be part of medical education. Like I knew going into it that’s part of the culture, and I just felt like this was just part of that.* -Participant B

Since mistreatment is expected, students felt that reports of mistreatment are not taken seriously. They had seen and heard of other students being mistreated, so mistreatment was not unique to them, and reporting was not truly warranted. For some, no action being taken on reports was an assumption, but for others, they had witnessed reports yielding no action by administrators.

> *I realize that I am in a hierarchical system, and anything I say is going to possibly backfire on me and make things even worse for me. So it kind of made me feel a little powerless.* -Participant D

Finally, some students noted that while they can report, actions taken by administrators would have little effect on the culture of mistreatment. They felt this was the true root of the problem, and thus felt reporting would be futile.

> *I think the reporting experience is adequate, but I don’t know like, if the culture will shift… Even if you have the information… the problems [that] exist are pervasive.* – Participant E

## Discussion

Medical education has a critically important hierarchy, meant to allow for the supervision of learners in a clinical environment while promoting the safe care of patients. This safety net allows for learners to check their clinical decision making with preceptors and allows for their personal growth. However, this study and others have shown how the hierarchy of medicine is not always protective. This hierarchy can perpetuate the cycle of student mistreatment and prevents reporting.

The peer-to-peer interview approach also allowed us to deeply explore individual episodes of mistreatment with the participants and understand their decision-making process. This allowed us to identify four main factors that influence reporting: personal, situational, structural, and climate. Within each factor, we sought to determine both factors that promoted reporting and prevented reporting. We found that only personal and situation factors promoted reporting and that students mostly reported reasons not to report. We also confirmed the large effect the culture and hierarchy of an institution can have a profound effect on whether a student chooses to report, and this can be exemplified through each factor.

Personally, the students struggle with possible repercussions on themselves and in the end, don’t trust themselves to report such information. The concern that reporting perpetrators within their intended specialty was consistent across other studies (Bell et al. 2021; Chung et al. 2018). These students sense the power that certain preceptors may hold over them and do not wish to take a risk. Unique to this study was the finding that participants wished to report due to a sense of justice. It seemed while there was not enough personal gain to report mistreatment, the collective gain to students was worth reporting mistreatment. Perhaps this is a motivating factor as the student may see reporting mistreatment as a means to change culture.

Situationally, participants struggle with the ambiguous definition of mistreatment, as has been shown in work by Gan and Snell (2014). In focus groups with students, Gan and Snell found that mistreatment that was subtle and common were more difficult to report. As a result, these instances were more distressing to the student. On the other end of the spectrum, participants found obvious instances of mistreatment easy to report.

Third party confidents or bystanders also impact reporting. Participants in this study went to a trusted advisor or family to disclose the event. Bell et al. (2021) described this phenomenon as “testing the waters.” Students use the reaction of confidants to determine whether to report, which can be a motivating factor. Medical students also use the reactions of bystanders in their reporting decision. Bystanders act as an informal opinion on the act of mistreatment, but also a visual representation of the medical hierarchy.

Strikingly, the bystanders described in this study varied from medical students to attendings, some of whom were the superior or peer of the perpetrator. Yet, none of the bystanders aided the students. Thus, the participants saw those around them not reacting, and perhaps felt that if no one came to their aid in the moment, then no one would believe or act on their reporting.

Structurally, students feel that a report will not be taken seriously, as they are at the bottom of the hierarchy. In a study of Australian medical students, the students lacked confidence that their institution would act (Colenbrander et al. 2020). Californian students in another study (Chung et al. 2018) described a feedback session with a course director where concerns about mistreatment were brushed off, thus the participants inferred that informing superiors of mistreatment would likely not yield results. Participants in this study and other studies noted the logistical barriers: not knowing where to report, not knowing where the information went, not wanting to spend the time reporting (Bell et al. 2021; Chung et al. 2018). Students noted their clinical demands were already extensive and further time spent on reporting would be a waste. These findings show that students find current reporting systems to be high effort and reporting is not worth the minimal gain.

Climate factors are the most transparent view of the hierarchy of medicine and its impact on students’ reporting. Participants in our study verbalized feeling that mistreatment was “expected” in the culture of medicine. They felt “powerless” in their pursuit of reporting due to the “hierarchical” nature of medicine. This was echoed in other studies of medical students in California, Canada, and Australia (Bell et al. 2021; Chung et al. 2018; Colenbrander et al. 2020). Students in other studies even called medical school a “hazing process,” (Chung et al. 2018) and they “kept their head down” (Bell et al. 2021) despite ongoing abuse. While reflecting on their experience and possible fixes, students in the present study were doubtful of improvement given a “culture shift” would be necessary for change. Thus, this study shows that the participants have awareness of the effect of the hierarchy of medicine on student mistreatment and their desire to report, and they feel powerless to change it.

For each factor, there are numerous mitigation strategies for an institution to consider. A robust reporting system grounded in improving the culture of an institution is critical. Institutions should publicize the reporting structure widely to allow trainees to readily utilize it when needed. Deidentified data regarding how reports are investigated, what actions are taken, and the number of reports per year should be published and accessible to students. The reporting system should also be easy to use, as medical students have been shown to weigh the amount of work it takes against their high clinical demands. There needs to be in place a vehicle for reporting retaliation that is anonymous and trusted by students to be anonymous. This may require institutions to completely overhaul their current reporting systems since it is our impression that most confidential reporting mechanisms are not experienced by students as confidential. Furthermore, if repercussions occur, the institutional response must be swift and firm since once retaliation for reporting, even one instance, occurs and is not addressed the entire reporting system is in jeopardy.

Finally, bystander training should be part of making a robust reporting system. Such training should focus on how bystanders can diffuse a situation and support those experiencing mistreatment, as previously implicated in the fight against sexual misconduct in medicine (Aggarwal and Brenner 2020). This training should be targeted at those higher in the medical hierarchy, as they will have the most power to intervene.

Outside of the reporting system, institutions need to take steps to ensure the culture is positive and supportive for trainees. This is best accomplished by a top-down approach, especially considering the findings of the impact of culture on reporting (Dankoski et al. 2014; Leape et al. 2012). Leadership in medical education sets the tone for how student mistreatment is viewed, handled, and tolerated. No intervention can be successful without the self-scrutiny and involvement of those involved in educational leadership. Given the well-established connection between learning environment and student empathy, improving the culture of an institution will likely lead to a better learning experience and create more empathic physicians (Brazeau et al. 2010; Neumann et al. 2011). Empathetic students become empathetic residents and preceptors, which can cease the cycle of mistreatment.

## Limitations

Medical student participants in our study sample all hailed from a single institution, which constituted a non-random sample of volunteers who self-selected to participate in the study. Our sample also had a higher incidence of reporting compared to the general population of medical students, which may limit the generalizability of our results. Lastly, nearly all of the incidents of mistreatment occurred in clinical settings; while student mistreatment in the pre-clinical setting may be less common, it does occur and may have different barriers to reporting than those identified in our study.

Future research should further develop the contribution of the mistreatment climate (i.e. a measure of a student’s perception of the culture of mistreatment within medical education) to willingness to report and the incidence of mistreatment. Within the safety literature, validated surveys have been created to assess safety climate, and this may be a fruitful avenue in the field of student mistreatment (Colla et al. 2005). Such an instrument may allow researchers to determine if interventions to improve the culture of an institution change the way trainees perceive the culture. Finally, the impact of witnesses on the reporting and identification of mistreatment should be more thoroughly researched.

## Conclusion

There are multiple factors students consider when deciding to report mistreatment, few of which are motivators to report. Unique to this study was participants sense of justice to report mistreatment and the negative impact of bystanders on reporting. Primarily targeting the culture of an institution may improve the reporting of student mistreatment, while also creating a positive learning environment and psychological safety for learners.

## Data Availability

All data produced in the present study are available upon reasonable request to the authors

## Acknowledgements

This manuscript was made possible by the Yale National Clinician Scholars Program. Grant support was provided to KB Montgomery by the Agency for Healthcare Research and Quality under grant T32 HS013852. Grant support was provided to Alissa Chen by NIH T32 AG019134.

## References

Aggarwal R, Brenner AM. 2020. #metoo: The role and power of bystanders (aka us). Academic Psychiatry. 44(1):5–10.

Ahmer S, Yousafzai AW, Bhutto N, Alam S, Sarangzai AK, Iqbal A. 2008. Bullying of medical students in pakistan: A cross-sectional questionnaire survey. PLoS One. 3(12):e3889.

Alzahrani HA. 2012. Bullying among medical students in a saudi medical school. BMC Res Notes. 5:335.

Bell A, Cavanagh A, Connelly CE, Walsh A, Vanstone M. 2021. Why do few medical students report their experiences of mistreatment to administration? Medical Education. 55(4):462–470.

Brazeau CM, Schroeder R, Rovi S, Boyd L. 2010. Relationships between medical student burnout, empathy, and professionalism climate. Acad Med. 85(10 Suppl):S33–36.

Byrne E, Brugha R, Clarke E, Lavelle A, McGarvey A. 2015. Peer interviewing in medical education research: Experiences and perceptions of student interviewers and interviewees. BMC Res Notes. 8:513.

Chung MP, Thang CK, Vermillion M, Fried JM, Uijtdehaage S. 2018. Exploring medical students’ barriers to reporting mistreatment during clerkships: A qualitative study. Med Educ Online. 23(1):1478170.

Colenbrander L, Causer L, Haire B. 2020. ’If you can’t make it, you’re not tough enough to do medicine’: A qualitative study of sydney-based medical students’ experiences of bullying and harassment in clinical settings. BMC Med Educ. 20(1):86.

Colla JB, Bracken AC, Kinney LM, Weeks WB. 2005. Measuring patient safety climate: A review of surveys. Qual Saf Health Care. 14(5):364–366.

Colleges AAoAM. 2023. Medical school graduation questionnaire: 2023 all schools summary report. Washington, D.C.

Cook AF, Arora VM, Rasinski KA, Curlin FA, Yoon JD. 2014. The prevalence of medical student mistreatment and its association with burnout. Acad Med. 89(5):749–754.

Dankoski ME, Bickel J, Gusic ME. 2014. Discussing the undiscussable with the powerful: Why and how faculty must learn to counteract organizational silence. Acad Med. 89(12):1610–1613.

Frank E, Carrera JS, Stratton T, Bickel J, Nora LM. 2006. Experiences of belittlement and harassment and their correlates among medical students in the united states: Longitudinal survey. BMJ. 333(7570):682.

Gágyor I. 2012. Frequency and perceived severity of negative experiences during medical education in germany--results of an online-survery of medical students. GMS Z Med Ausbild. 29(4):1–12.

Gan R, Snell L. 2014. When the learning environment is suboptimal: Exploring medical students’ perceptions of “mistreatment”. Acad Med. 89(4):608–617.

Haglund ME, aan het Rot M, Cooper NS, Nestadt PS, Muller D, Southwick SM, Charney DS. 2009. Resilience in the third year of medical school: A prospective study of the associations between stressful events occurring during clinical rotations and student well-being. Acad Med. 84(2):258–268.

Hariharan TS, Griffin B. 2019. A review of the factors related to burnout at the early-career stage of medicine. Med Teach. 41(12):1380–1391.

Haviland MG, Yamagata H, Werner LS, Zhang K, Dial TH, Sonne JL. 2011. Student mistreatment in medical school and planning a career in academic medicine. Teach Learn Med. 23(3):231–237.

Heru A, Gagne G, Strong D. 2009. Medical student mistreatment results in symptoms of posttraumatic stress. Academic Psychiatry. 33(4):302–306.

Hockey J. 1993. Research methods -- researching peers and familiar settings. Research Papers in Education. 8(2):199–225.

Leape LL, Shore MF, Dienstag JL, Mayer RJ, Edgman-Levitan S, Meyer GS, Healy GB. 2012. Perspective: A culture of respect, part 2: Creating a culture of respect. Acad Med. 87(7):853–858.

Mavis B, Sousa A, Lipscomb W, Rappley MD. 2014. Learning about medical student mistreatment from responses to the medical school graduation questionnaire. Acad Med. 89(5):705–711.

Mearns KJ, Flin R. 1999. Assessing the state of organizational safety—culture or climate? Current Psychology. 18(1):5–17.

Moscarello R, Margittai KJ, Rossi M. 1994. Differences in abuse reported by female and male canadian medical students. CMAJ. 150(3):357–363.

Neumann M, Edelhauser F, Tauschel D, Fischer MR, Wirtz M, Woopen C, Haramati A, Scheffer C. 2011. Empathy decline and its reasons: A systematic review of studies with medical students and residents. Acad Med. 86(8):996–1009.

Patton MQ. 2002. Qualitative research & evaluation methods. Thousand Oaks, CA: Sage Publications.

Richman JA, Flaherty JA, Rospenda KM, Christensen ML. 1992. Mental health consequences and correlates of reported medical student abuse. JAMA. 267(5):692–694.

Silver HK, Glicken AD. 1990. Medical student abuse: Incidence, severity, and significance. JAMA. 263(4):527–532.

